# Development and Validation of The Activity-Based Checks (ABCs) of Pain: A Functional Pain Scale

**DOI:** 10.1101/2024.01.29.24301629

**Authors:** Celina G. Virgen, Robert Wright, Bryan Renslo, Tuleen Sawaf, Hanna Moradi, Maria Edelen, Jennifer Villwock

## Abstract

The Activity-Based-Checks of Pain (ABCs) is a pain assessment tool incorporating activities of daily living and instrumental activities of daily living. Unlike widely used pain scales which are oftentimes unidimensional and highly subjective, the ABCs was designed to focus on function capabilities and limitations of patients due to pain. This study sought out to validate the factorial structure of the ABCs and assess its use in participants with chronic pain. Participants were recruited in two phases from Prolific – an online service designed to identify research participant recruitment based on study criteria. Phase one optimized the design of the ABCs, with 297 subjects selecting their preferred icon for each function and rating its understandability. The most preferred and understandable icons were then used in phase two, where 304 chronic pain participants completed the ABCs, PROMIS-29, additional PROMIS items that were analogous to the ABCs functions but not represented in the PROMIS-29, and the Brief Pain Inventory (BPI). Data was analyzed using exploratory factor analysis and confirmatory factor analysis demonstrating four factor loadings: multi-planal activities, sitting/hip flexor pain, walking/ambulation, and pain interference with lightweight unilateral activities. High internal consistency was demonstrated with all four factor loadings. Correlations between items in the ABCs, PROMIS, and BPI resulted in moderate to strong correlations demonstrating strong evidence for the validity of the ABCs as a functional pain assessment tool.

## Introduction

Pain is a unique experience that is personal, private, subjective, and can become a barrier to achieving an enjoyable quality of life [4]. Assessing pain can provide insight into the type of pain experienced by patients, if current treatment is adequate for pain control, or if additional interventions are merited [8]. Despite a number of pain assessment tools, poor pain assessment remains one of the major barriers to healthcare professionals appropriately treating pain [7].

There is an inherent disconnect between patient-clinician communication regarding pain intensity and management goals [11]. Patients have expressed preference and trust in clinicians who go beyond focusing on the physical pain [10]. Current tools limit the healthcare provider in obtaining objective measures and patient pain experience. Tools such as the Numeric Rating Scale (NRS) and Wong-Baker Faces Pain Rating Scale (FACES) are highly subjective which may misrepresent pain intensity score and tolerable pain while overshadowing function and patient progress [1,25]. Additionally, studies have suggested that recorded pain levels do not reliably predict subsequent treatment [12,26]. Widely accepted scales are unidimensional; error may be introduced when attempting to gauge the intensity of pain at different points in time, during activities, and movements [4,12]. The complexity of the pain experience is even greater for patients with chronic pain because so many aspects and activities of their lives are impacted [4].

Due to the multifaceted nature of chronic pain, adequately capturing pain’s impact on a patient’s life is challenging. Simple numeric rating scales lack the necessary nuance. More detailed, but lengthy, Likert-style questionnaires require significant cognitive energy to both complete and interpret. The Activity-Based-Checks of Pain – Functional Pain Scale (ABCs) is a visual/infographic pain assessment that was created to circumvent these issues. It uses simple visual icons to represent activities and is comprised of various activities of daily living (ADL), instrumental activities of daily living (IADL), and other activities a patient may encounter in their daily life. This scale is meant to not only evaluate pain but also understand how pain interferes and limits activities. The objectives of this study are to (1) investigate and validate the factorial structure of the ABCs and (2) evaluate its use in chronic pain patients.

## Methods

Institutional Review Board and ethical approval for this study was obtained from the University of Kansas Medical Center, study 00142379.

### Target population and Recruitment

Participants were identified and recruited via Prolific, an online source used to identify volunteer research participants based on study criteria [23]. Two separate cohorts were recruited. The first cohort comprised a representative sample of United States adults and was used to determine the best-fit icons for each activity or function described in terms of icon comprehensibility and strength-of-association. The best-fit icons were incorporated into the next iteration of the ABCs. Arrow style to indicate where pain level was on the pain spectrum was also tested with subjects selecting between a black arrow, a colorful arrow, and arrows annotated with descriptors of mild, moderate, or severe pain. Field testing of the ABCs was done in the second cohort, who had all experienced chronic pain for at least 6 months. Participants in both cohorts were recruited without restrictions of race or sex, were age 18+, resided in the United States, and used English as their primary language.

### Content Validity – Cohort 1

Participants were presented with the name of the function or activity of interest (e.g. preparing a meal) and four icons meant to represent it. They selected their preferred icon based on relevance to the function (In your opinion, which of these images represents preparing a meal?), comprehensibility (Is it easy or difficult to understand that these images represent preparing a meal?), and strength-of-association (Which of the images best represents the ability to prepare a meal?). The most-selected and best-fit icon was then used in the ABCs presented to cohort two (chronic pain participants). Additionally, participants were able to provide their own opinion of a potential icon for such activity or function listed.

### ABC’s of Pain Functional Scale

Initial development of the ABCs included selection of icons representative of the activity or functions: walking, jogging/running, prolonged standing, prolonged sitting, sleeping, getting out of bed, sitting up, getting out of a chair, doing laundry, grocery shopping, walking a dog, playing with a child, bathing, housework, picking up 20 lbs, preparing a meal, eating, opening bottle to take medicine, walking upstairs, talking on the phone, getting dressed, personal care/grooming, driving, and toileting. Completion of the ABCs was done in a stepwise form. First, participants were asked if they are able to perform the given activity and select “yes”, “no” or “not applicable” as an option (Image 1). If the participant responded “yes” to such activity, then they were able to indicate their level of pain on a bidirectional arrow from “no pain” to “worst pain”. If they selected “no”, a follow-up question asks why and participants may select that the activity is too painful or for other reasons. If “too painful”, then participants were able to indicate on the bidirectional arrow how much pain they thought they would be in if the activity was attempted. The bidirectional arrow was scored with 0 being “no pain” to 10 being “worst pain”.

**Image 1.**
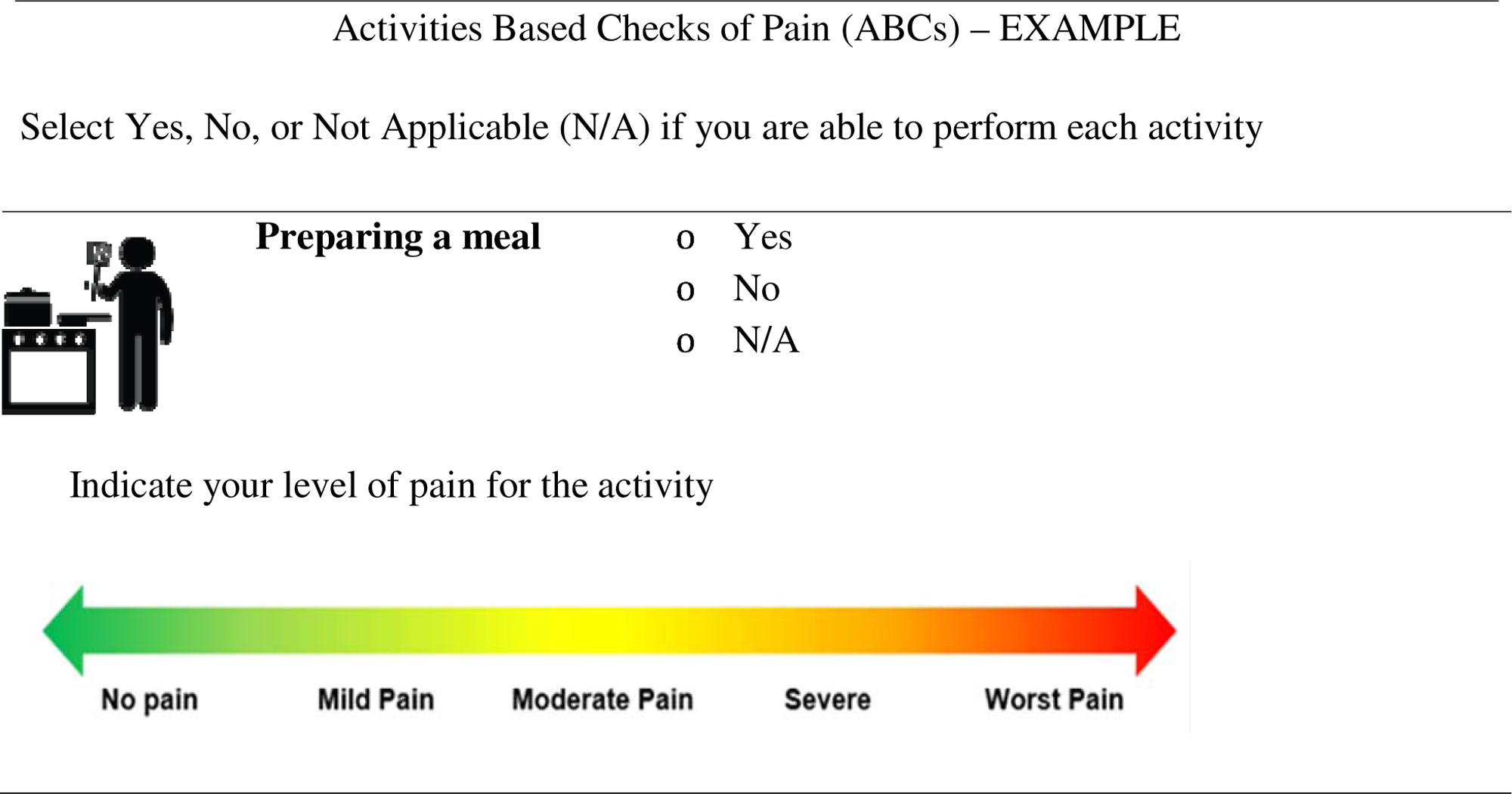
Activities Based Checks of Pain – Example.

### Cohort 2 - Construct Validity

Participants were recruited to evaluate construct validity of the ABCs against the Brief Pain Inventory (BPI) and the Patient Reported Outcomes Measurement Information System (PROMIS)-29, both of which are previously validated measures of pain. The BPI is meant to provide insight on the severity of pain experienced and the effect pain has on daily functioning in multiple domains as previously described [3]. Higher scores indicate worse pain and pain interference. Items in the BPI regarding everyday pain and percentage of pain relief with treatment are optional and are not included in the scoring. A score of 1 - 4 is considered overall mild pain, 5 - 6 is moderate pain, while a score average of 7 - 10 is severe pain. The PROMIS-29 was developed to measure various health aspects unrelated to any specific disease such as self-efficacy, fatigue, anxiety, depression, pain interference, sleep, and physical function [5]. Responses are on a 5-point Likert scale and scored from 1 - 5. The PROMIS-29 also includes a numeric rating scale which is scored from 0 - 10. A total score is based on the sum of all items producing a score between 4 - 20 which is then converted into a T-score with a mean of 50 and standard deviation of 10. The ABCs contained more functional items than the PROMIS-29. As such, the PROMIS item bank was searched so that items analogous to all the ABCs functions were included so that comparisons between ABCs and PROMIS items could be made. Twenty-three items were added related to pain inference and physical function making the PROMIS a total of 52 questions. Time necessary to complete the BPI, PROMIS-29 plus additional items, and ABCs were also recorded.

Subjects were given the opportunity to provide open-ended narrative feedback on their experience with the different pain scales as well as salient experiences they had with the health care system as people with chronic pain. To assess for test-retest reliability, cohort two participants were invited to complete pain assessments two weeks after their initial participation.

### Statistical Analysis

All study data were collected and managed using REDCap electronic data capture tools and exported to R (4.2.1) for analysis.

Responses from Cohort 1 were simply majority voting regarding the most understandable and representative item and arrow to use in the final version of the ABCs.

To evaluate the construct validity of the ABCs, we conducted an exploratory factor analysis (EFA) to examine the structure underlying responses to the items. The factor analysis results were used to determine the number of latent factors and their relationship to theory. We conducted a maximum likelihood analysis with oblique rotation. The appropriate number of factors was determined based on the number of eigenvalues greater than one, the scree plot, parallel analysis, and the interpretability of the factors. The factor loadings were examined to identify items that loaded on each factor.

Confirmatory factor analysis (CFA) was conducted to confirm the factor structure identified through EFA. The analysis was conducted using maximum likelihood estimation to account for possible correlation amongst the factor components. Model fit was assessed using the chi-square test, comparative fit index (CFI), Tucker-Lewis index (TLI), and root mean square error of approximation (RMSEA). The chi-square statistic is very sensitive to sample size and frequently rejects valid models due to large samples (n > 100). Modification indices were examined to identify areas of misfit and to refine the model.

Internal consistency was assessed using Cronbach’s alpha coefficient for each factor identified through EFA and confirmed with CFA. A coefficient alpha of ≥ 0.70 was considered acceptable [24]. Test-retest reliability was assessed using the intraclass correlation coefficient (ICC). A minimum acceptable value for intraclass correlations for multi-item scales was set at ≥ 0.70 [17].

Convergent validity was assessed by examining the correlation between the new pain scale and other measures of pain intensity. Pearson’s correlation coefficient was used to assess the correlation between the ABCs, BPI, and PROMIS. The strength of the correlation will be interpreted using the following guidelines: r < 0.3 = weak correlation, r = 0.3-0.6 = moderate correlation, r > 0.7 = strong correlation [2].

## Results

### Participant Demographics

Electronic online consent was provided and collected for 297 participants in cohort one and 304 in cohort two. In cohort one, 55% were females with 70.1% identifying as White with a mean age of 34. Participants in cohort two were 51% male and 80% White with a mean age of 41 years with over half reporting chronic pain issues of at least 5 years or more. (Table 1). Table 2 details descriptive statistics for the ABCs of pain subscales along with the PROMIS-29 pain interference, BPI pain interference, SF-20 pain severity, and the NRS.

**Table 1.** Participant Characteristics.

| Characteristic | Icon selection<br>participants<br>N = 297<sup>1</sup> | All participants<br>N = 304<sup>1</sup> | Re-test participants<br>N = 194<sup>1</sup> |
| --- | --- | --- | --- |
| Age | 34 (12) | 41 (14) | 43 (14) |
| Sex |  |  |  |
| Female | 164 (55%) | 144 (48%) | 95 (51%) |
| Male | 132 (44%) | 152 (51%) | 90 (49%) |
| Prefer not to say | 1 (0.3%) | 2 (0.7%) | 0 (0%) |
| Race |  |  |  |
| Asian | 18 (6.1%) | 10 (3.4%) | 6 (3.2%) |
| African American/Black | 26 (8.8%) | 19 (6.4%) | 11 (5.9%) |
| White | 210 (70.7%) | 238 (80%) | 148 (80%) |
| Mixed | 29 (9.8%) | 21 (7.0%) | 13 (7.0%) |
| Other | 13 (4.4%) | 10 (3.4%) | 7 (3.8%) |
| Pain Duration |  |  |  |
| 6-12 months | - | 22 (7.4%) | 10 (5.4%) |
| 1-2 years | - | 16 (5.4%) | 9 (4.9%) |
| 2-5 years | - | 76 (26%) | 47 (25%) |
| 5-10 years | - | 106 (36%) | 68 (37%) |
| 10-20 years | - | 78 (26%) | 51 (28%) |

<sup>1</sup> Mean (SD); n (%)

**Table 2.**
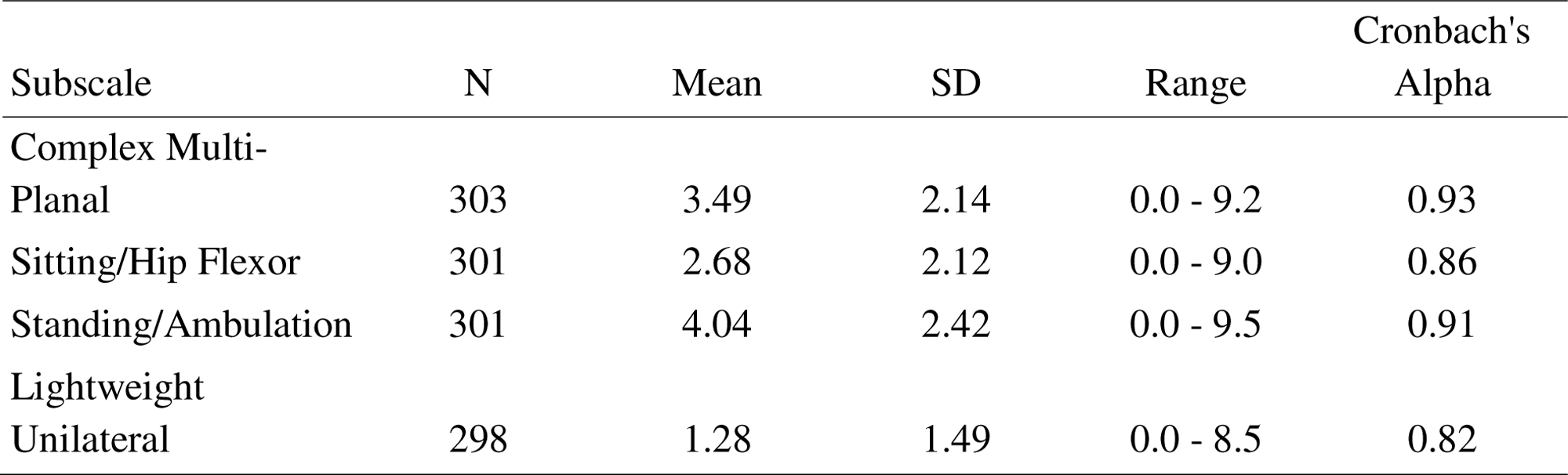
Descriptive statistics and internal consistency reliability for ABCs of Pain subscales.

The score distribution and pattern response for the ABCs of each activity is reported in Table 3. “Talking on the phone” and “Opening a bottle of medicine” show evidence of a floor effect with 62% and 57% respectively selecting “0” while there was no ceiling effect for any items. The average time to take the BPI, PROMIS-29, and ABCs was 2.4 minutes, 3.8 minutes, and 3.9 minutes respectively.

**Table 3.** Score distributions, response frequencies and averages for the ABCs.

| Activities/Functions | 0 | 1 | 2 | 3 | 4 | 5 | 6 | 7 | 8 | 9 | 10 | Mean (SD) |
| --- | --- | --- | --- | --- | --- | --- | --- | --- | --- | --- | --- | --- |
| Walking | 43 | 43 | 38 | 48 | 41 | 13 | 31 | 20 | 10 | 3 | 2 | 3.24 (2.44) |
| Jogging/Running | 21 | 20 | 20 | 30 | 19 | 22 | 53 | 28 | 29 | 12 | 21 | 5.08 (2.91) |
| Sleeping | 53 | 50 | 39 | 52 | 25 | 26 | 24 | 13 | 6 | 2 | 3 | 2.88 (2.37) |
| Getting out of bed | 62 | 54 | 55 | 45 | 26 | 12 | 17 | 15 | 6 | 3 | 0 | 2.53 (2.25) |
| Sitting up | 76 | 62 | 43 | 41 | 24 | 16 | 17 | 9 | 3 | 4 | 0 | 2.28 (2.2) |
| Doing laundry | 62 | 50 | 53 | 39 | 28 | 20 | 18 | 10 | 6 | 3 | 0 | 2.55 (2.24) |
| Grocery shopping | 51 | 39 | 54 | 41 | 30 | 21 | 26 | 16 | 12 | 3 | 1 | 3.05 (2.43) |
| Walking a dog | 30 | 21 | 32 | 41 | 21 | 22 | 14 | 12 | 10 | 3 | 4 | 3.41 (2.52) |
| Playing with a child | 19 | 30 | 33 | 23 | 24 | 19 | 23 | 17 | 7 | 2 | 4 | 3.62 (2.52) |
| Bathing | 80 | 57 | 47 | 46 | 25 | 19 | 12 | 8 | 5 | 0 | 0 | 2.18 (2.05) |
| Housework | 26 | 30 | 42 | 49 | 32 | 28 | 33 | 32 | 17 | 9 | 1 | 3.94 (2.51) |
| Picking up 20 lbs | 15 | 33 | 32 | 31 | 30 | 41 | 34 | 37 | 15 | 11 | 9 | 4.47 (2.63) |
| Preparing a meal | 63 | 52 | 51 | 49 | 22 | 15 | 19 | 13 | 7 | 0 | 0 | 2.49 (2.19) |
| Walking up stairs | 39 | 51 | 41 | 43 | 19 | 23 | 14 | 25 | 18 | 10 | 4 | 3.44 (2.76) |
| Driving | 57 | 51 | 48 | 45 | 14 | 19 | 9 | 12 | 6 | 1 | 0 | 2.40 (2.16) |
| Toileting | 109 | 67 | 46 | 22 | 17 | 18 | 6 | 6 | 4 | 1 | 0 | 1.7 (1.99) |
| Prolonged standing | 31 | 22 | 28 | 44 | 27 | 18 | 34 | 40 | 24 | 14 | 7 | 4.43 (2.82) |
| Prolonged sitting | 66 | 37 | 43 | 35 | 15 | 23 | 33 | 22 | 10 | 2 | 5 | 3.1 (2.69) |
| Getting out of a chair | 61 | 55 | 47 | 48 | 23 | 17 | 25 | 15 | 6 | 0 | 0 | 2.6 (2.23) |
| Talking on the phone | 177 | 44 | 27 | 15 | 8 | 5 | 3 | 2 | 3 | 1 | 1 | 0.96 (1.73) |
| Eating | 130 | 68 | 33 | 30 | 10 | 10 | 4 | 6 | 3 | 0 | 1 | 1.4 (1.87) |
| Opening a bottle of medicine | 166 | 52 | 23 | 21 | 13 | 6 | 3 | 0 | 4 | 0 | 1 | 1.05 (1.71) |
| Getting dressed | 91 | 70 | 41 | 34 | 20 | 18 | 12 | 5 | 6 | 2 | 0 | 2.0 (2.12) |
| Personal care/Grooming | 114 | 66 | 41 | 38 | 13 | 11 | 5 | 5 | 1 | 2 | 0 | 1.55 (1.83) |

### Factor Structure and Analysis

EFA was conducted to examine the underlying structure of the ABCs. A maximum likelihood analysis with oblique rotation was used. Oblique rotations were used with the hypothesis that different factors of pain would be correlated with each other. The factor analysis revealed a four-factor solution, explaining 69% of the total variance. The first factor consisted of six items and had loadings of 0.623 to 0.879 for items 1 to 6 with 24% of the variance explained. The second factor consisted of four items and had loadings of 0.577 to 0.824 for items 6 to 10 with 17% of the variance explained. The third factor consisted of four items and had loadings of 0.512 to 0.841 for items 11 to 14 with 16% of the variance explained. The fourth factor consisted of four items and had loadings of 0.519 to 0.817 from items 15 to 18 with 13% of the variance explained. The factor loadings suggest that the first factor measures pain interference with complex multi-planal activities. The second factor measures pain interference with sitting/hip flexor pain. The third is pain interference with walking/ambulation and the fourth factor is pain interference with lightweight unilateral activities.

CFA was conducted to confirm the factor structure identified through EFA (Table 4). The results of the CFA supported the four-factor structure. The chi-square test χ^2^ = 318.8, df = 129, p = <0.001. The comparative fit index (CFI) was 0.93, the Tucker-Lewis index (TLI) was 0.91, and the root mean square error of approximation (RMSEA) was 0.08, which all indicate a moderate fit.

**Table 4.** Final CFA of the ABCs scales.

| CFI | TLI | RMSEA | SRMR |
| --- | --- | --- | --- |
| 0.92 | 0.91 | 0.09 | 0.05 |

### Reliability

Internal consistency was assessed using Cronbach’s alpha coefficient for each factor identified through EFA and CFA. The alpha coefficients for pain interference with complex multi-planal activities, sitting/hip flexor pain, walking/ambulation and lightweight unilateral activities were 0.93, 0.86, 0.91, and 0.82 respectively, indicating high internal consistency. All participants who initially completed the survey were invited for test-retest at two weeks after initial responses. As shown in Table 1, 194 participants completed all assessments. Test-retest reliability was assessed using the intraclass correlation coefficient (ICC). The ICC for the total scale was 0.74, indicating moderate test-retest reliability.

### Convergent Validity

Convergent validity was assessed by examining the correlation between the ABCs subscales and other measures of pain interference (Table 5). Correlations between analogous items from pain scales are shown on Table 6. All correlations were found to be moderate to strong.

**Table 5.** ABCs subscale correlations with BPI, PROMIS-29 and NRS.

Tables 5. ABCs subscale correlations with BPI, PROMIS-29 and NRS
| ABCs of Pain Subscales |  |  |  |  |
| --- | --- | --- | --- | --- |
|  | Complex Multi-Planal | Sitting/Hip Flexor | Standing/Ambulation | Lightweight Unilateral |
| NRS | 0.43 | 0.39 | 0.41 | 0.37 |
| SF-20 (pain severity) | 0.59 | 0.50 | 0.59 | 0.37 |
| BPI (pain interference) | 0.58 | 0.51 | 0.59 | 0.48 |
| PROMIS-29 (pain interference) | 0.70 | 0.52 | 0.60 | 0.49 |

*Note: all values were significant ( $P < 0.001$ )*

**Table 6.**
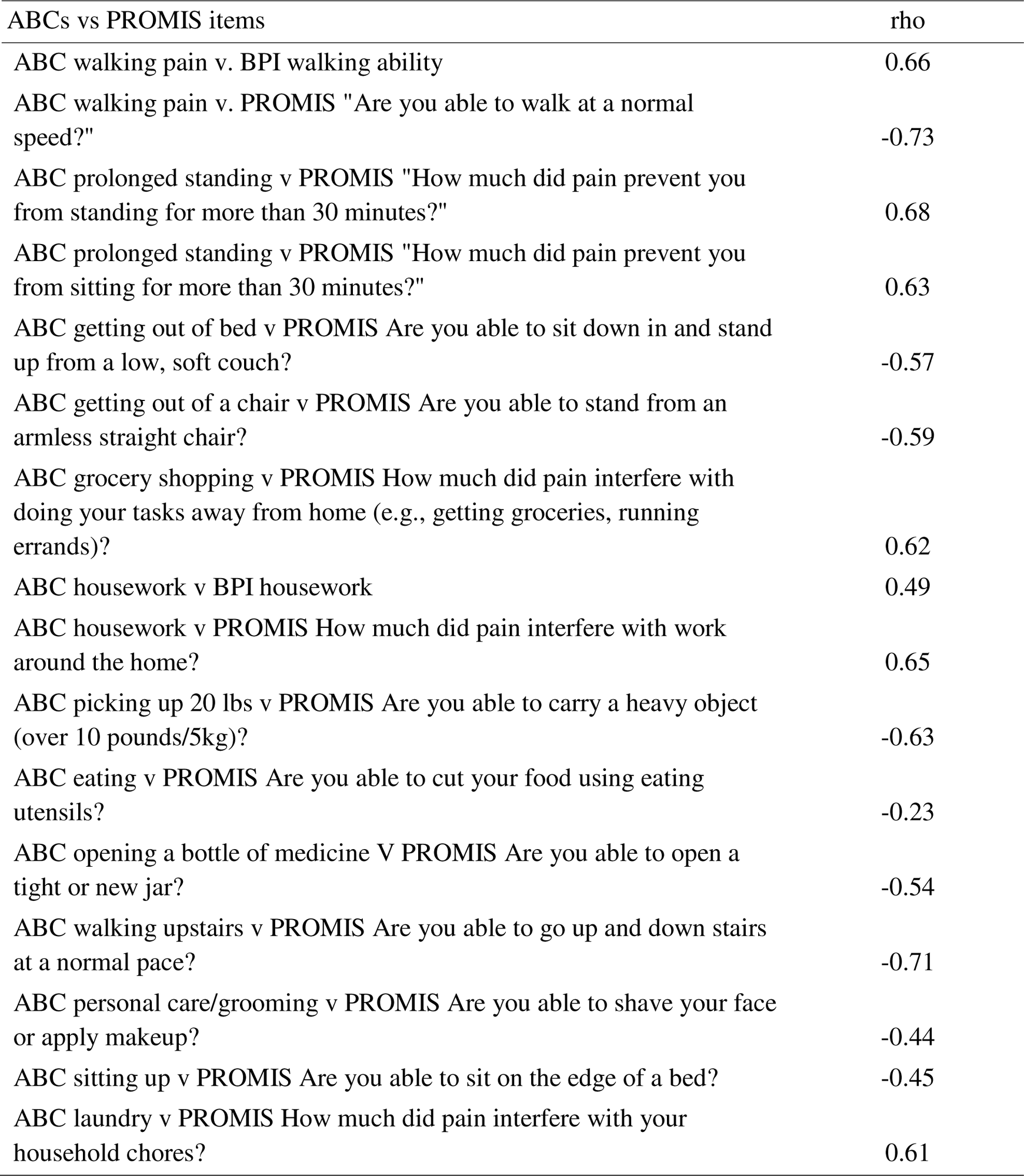
Spearman’s correlation coefficients for analogous variables.

| ABCs vs PROMIS items | rho |
| --- | --- |
| ABC walking pain v. BPI walking ability | 0.66 |
| ABC walking pain v. PROMIS "Are you able to walk at a normal speed?" | -0.73 |
| ABC prolonged standing v PROMIS "How much did pain prevent you from standing for more than 30 minutes?" | 0.68 |
| ABC prolonged standing v PROMIS "How much did pain prevent you from sitting for more than 30 minutes?" | 0.63 |
| ABC getting out of bed v PROMIS Are you able to sit down in and stand up from a low, soft couch? | -0.57 |
| ABC getting out of a chair v PROMIS Are you able to stand from an armless straight chair? | -0.59 |
| ABC grocery shopping v PROMIS How much did pain interfere with doing your tasks away from home (e.g., getting groceries, running errands)? | 0.62 |
| ABC housework v BPI housework | 0.49 |
| ABC housework v PROMIS How much did pain interfere with work around the home? | 0.65 |
| ABC picking up 20 lbs v PROMIS Are you able to carry a heavy object (over 10 pounds/5kg)? | -0.63 |
| ABC eating v PROMIS Are you able to cut your food using eating utensils? | -0.23 |
| ABC opening a bottle of medicine V PROMIS Are you able to open a tight or new jar? | -0.54 |
| ABC walking upstairs v PROMIS Are you able to go up and down stairs at a normal pace? | -0.71 |
| ABC personal care/grooming v PROMIS Are you able to shave your face or apply makeup? | -0.44 |
| ABC sitting up v PROMIS Are you able to sit on the edge of a bed? | -0.45 |
| ABC laundry v PROMIS How much did pain interfere with your household chores? | 0.61 |

**Table 7.** Maximum likelihood factor bases loadings for the retained 18 items of the ABCs.

| Activities/Functions | Four factor loadings |  |  |  |
| --- | --- | --- | --- | --- |
|  | Multi-planal movement | Sitting/Hip flexor | Walking/Ambulation | Light weight unilateral |
| Walking | 0.219 | - | 0.714 | - |
| Jogging/Running | - | 0.112 | 0.841 | - |
| Getting out of bed | 0.191 | 0.577 | - | - |
| Sitting up | - | 0.824 | - | - |
| Doing laundry | 0.704 | 0.231 | - | - |
| Grocery shopping | 0.879 | - | 0.131 | - |
| Walking a dog | 0.692 | - | 0.248 | - |
| Housework | 0.283 | - | 0.512 | - |
| Picking up 20 lbs. | 0.732 | 0.127 | - | - |
| Preparing a meal | 0.623 | 0.19 | - | - |
| Walking up stairs | 0.643 | - | - | 0.132 |
| Prolonged standing | - | - | 0.739 | 0.147 |
| Prolonged sitting | - | 0.748 | - | - |
| Getting out of a chair | - | 0.738 | 0.152 | - |
| Talking on the phone | - | -0.109 | - | 0.817 |
| Eating | - | 0.239 | - | 0.519 |
| Opening a bottle of medicine | -0.143 | - | 0.126 | 0.771 |
| Personal care/Grooming | 0.267 | - | - | 0.707 |

### Narrative Feedback on Pain Assessment

All participants had the opportunity to provide feedback on the ABCs as compared to the BPI and PROMIS. Comments from participants that indicated preference towards the ABCs, were: *“[ABCs] Makes me feel like I am not comparing myself to others”, “More rounded out picture of how my pain is limiting my life… then simply saying it’s an 8”, “My 4 may be someone else’s 7”,* and *“it gives the best idea of what my day to day is like”.* Participants commonly mentioned being able to communicate more detailed information about their pain especially when discussing with a healthcare provider. For participants that did not prefer the ABCs, comments included *“[0-10 scale] is easier to follow”, “Other scales give a definite number”*, and *“The* [0–10] *scale is what I grew up with and has always gotten the point across for me”*.

## Discussion

The aim of this study was to investigate and validate the factorial structure of the ABCs – a novel functional pain scale that utilizes a visual/icon-based format and assesses pain in the context of its impact on ability to perform activities of interest – and evaluate its use in chronic pain patients. Our results indicate the scale functions well at measuring pain interference. Our multidimensional scale measures pain interference with (1) complex multi-planal activities, (2) sitting/hip flexor pain, (3) walking/ambulation, and (4) lightweight unilateral activities. Test-retest reliability was moderate to strong with an ICC of 0.74. Fit indices from the CFA support the construct validity and the Cronbach’s alpha supports the internal consistency of the ABCs. The results confirm the use of the ABCs in chronic pain patients as a valid and reliable tool for assessing pain’s impact on function.

Confirmatory factor analysis with four-factor structure supported the validity of the ABCs. Among the four factors, similarities were noted creating the constructs of complex multi-planal activities, sitting/hip flexor pain, walking/ambulation, and lightweight unilateral activities. There were cross loadings for items such as “Walking a dog” with the walking/ambulation group of items. While this intuitively makes sense the item remained within the multiplanar subscale since walking a dog is an activity that will involve more than simply walking. Similarly, we see housework, getting out of bed, eating, and personal care/grooming show similar signs of cross loading onto factors that incorporate some of the aspects of the activity.

In the current study, the ABCs were found to correlate with the BPI and PROMIS scales demonstrating high external validity. Previous studies comparing results between the BPI and PROMIS when assessing pain have found variable results. Kean et al., evaluated persistent musculoskeletal pain in 250 patients before and after interventions using the BPI, PROMIS pain interference measures, the 2-item Bodily Pain subscale of the Medical Outcomes Study SF-36 bodily pain subscale, and the 3-item Pain Intensity, Enjoyment of Life, General Activity (PEG) Scale [15]. Compared to the BPI, the PROMIS was less sensitive in re-evaluating pain and detecting changes or improvements after intervention. In contrast, Chen et al., found no significant difference in 759 patients between the BPI and PROMIS in being able to assess pain at baseline and at 3 – 6 month follow-up [6]. Variability in responses to pain assessment can be highly context specific – for example, complex nature of pain, pain location, personal perceptions of pain, timing of pain evaluations – which may explain both consistency in results and lack thereof depending on study design [4,16]. In spite of this, the ABCs were able to capture tolerable and intolerable activities as reported by participants while having a moderately strong test re-test reliability. The ability of the ABCs scale to assess pain with activities may add value for physicians and health care team members who are trying to assess specific functional outcomes related to pain management interventions.

The ABCs employs a straightforward 0-10 pain scale as a heuristic, making it easily comprehensible and applicable for both practitioners and patients. It builds on this concept with subscales created from items grouped based on similar movements and body parts, thus allowing for more nuance when evaluating pain. The capturing of this more nuanced data can help facilitate better communication of pain between physicians and patients as well as among pain management teams. Tracking and understanding pain with specific activities can help with understanding and improving pain treatments and interventions.

Narrative feedback from participants highlighted several positive aspects of the ABCs, most notably its ability to provide comprehensive information about their lived experience and reducing pain’s subjectivity. While the ABCs did not appear to be the best tool for all participants, many preferred a detailed personalized scale to describe their experience with pain. In a study by van Dijk et al., inconsistencies regarding interpretation of a reported score on the commonly used 0-10 Numeric Rating Scale (NRS) were found [25]. The same numeric score did not mean the same thing to different patients. Depending on the individual, reporting a pain score of 4 could be described as “bearable” or “unbearable.” This was also the case for a 6 on the NRS. In a study investigating pain sensitivity and lower back pain, the threshold for pain did not equate to disability or functional impact [13]. Miscommunication of pain may be further influenced by bias, the perception of pain by the healthcare provider, and cultural factors that determine what a patient feels is appropriate to share with their care teams [20,21]. For example, in some cultures, discussing pain may be considered to be inappropriate complaining or rude due to an implication that the pain is related to provision of poor care [18,19]. A personalized pain scale like the ABCs that is linked to the objective metric of being able to perform an activity, may be a useful tool to reduce biases and inconsistencies in pain communication for chronic pain patients.

The ABCs is intended to provide meaningful information that will visually and efficiently reflect pain and the impact on daily function. When comparing time to completion among the three scales, the ABCs completion time was similar to the PROMIS items, 3.9 minutes vs 3.8 minutes, respectively. However, compared to the BPI (2.4 minutes), the ABCs was approximately 1.5 minutes longer to complete. A future intent of the ABCs is to create an adaptable scale that is personalized to each patient’s functional priorities. Not every activity or function in the current ABCs scale is applicable to all patients. The PROMIS has become a well-recognized scale with numerous bank items that can be added or removed from the scale based on clinician preference and purpose for its use. Time to completion for adaptations of the PROMIS have been previously reported to range from 3.3 minutes to 6.4 minutes [14,22]. In comparison, the BPI has been reported to take up to 5 minutes [9]. Time to completion for the BPI and PROMIS varies depending on the target population and the version of the scales being implemented. Future adaptations of the ABCs, with the ability to select pertinent activities, may further decrease time to completion.

### Limitations

Although the results of the study indicate validity of the ABCs, the study was not without limitations. First, participants were recruited to participate via a survey distribution site based on self-reported chronic pain. This limited researchers from confirming chronic pain as a medical diagnosis. Second, test-retest was conducted, however, given the complexity of pain and individual lifestyles, levels of pain and location may fluctuate. Despite this, test-retest results do not appear to have been affected. We acknowledge that there is variability within the chronic pain patient population and that included subjects could have chronic pain from differing etiologies. However, we do not believe this to have significantly influenced our results, particularly in comparison between assessments as each individual served as their own control. While the four subscales cover a broad range of activities, there are certainly more activities that could be included in the ABCs depending on priorities of a particular patient population or recovery following certain procedures. Additional studies will address such patient- and procedure-specific outcomes.

## Conclusion

This study successfully validated the ABCs, a novel functional pain scale that employs a visual/icon-based format to assess pain interference in chronic pain patients. The scale’s multidimensional structure, as identified through EFA and confirmed through CFA, measures pain interference in four distinct areas: complex multi-planar activities, sitting/hip flexor pain, walking/ambulation, and lightweight unilateral activities. The ABCs demonstrated moderate to strong test-retest reliability and fit indices from the CFA supported its construct validity and internal consistency. The scale’s correlation with the BPI and PROMIS scales further supports its external validity. The ABCs offers a nuanced and functional approach to evaluating pain with evidence for its validity in chronic pain patients.

## Data Availability

All data produced in the present study are available upon reasonable request to the authors

## Acknowledgements

This project was funded by the National Institute of Health as a part of the National Center for Advancing Translational Sciences KL2 Mentored Clinical Research Scholars Award, Parent award UL1TR002366. The authors have no conflict of interest.

